# Tortuosity of the iliac artery is associated with ipsilateral low back pain – a case-control study

**DOI:** 10.1101/2025.02.28.25322883

**Authors:** Jens Maier

## Abstract

**Background:** Thrust in arterial bends causes enhanced pulsatility.

**Purpose:** To study the association between tortuous iliac arteries and lateral low back pain, hypothesizing that pulsatile microtrauma is inflicted on the psoas muscle.

**Materials and methods:** This retrospective case-control study compared the self-reported frequency and localization of low back pain between subjects with normal (control group; n=31; mean age 62.5y) and markedly tortuous (study group; n=31; mean age 63.2y) iliac arteries. The groups consisted of 17 male and 14 female age-matched pairs. All participants were outpatients aged 40 to 80 y having undergone abdominal computed tomography (CT) without significant abnormalities. They completed a questionnaire on low back pain. Marked iliac tortuosity, defined as arterial bends of 90 deg. or sharper, was subdivided into bends targeting the psoas muscle and bends passing in front of or behind the psoas muscle.

**Results:** Lateral low back pain on one or both sides was reported more often in the study group (n=14) than in the control group (n=6), p=0.031. Lateral low back pain was reported on 11 of 20 sides of a markedly tortuous iliac artery targeting the psoas muscle, but only on 8 of 62 sides in the control group, p=0.0003. In subjects with marked iliac tortuosity targeting the psoas muscle on one side, the side of targeting correlated with the side of pain, p=0.032.

**Conclusion:** Iliac artery tortuosity targeting the psoas muscle is associated with ipsilateral low back pain. This association may explain low back pain in some of the cases currently regarded as nonspecific.

## Introduction

Low back pain is highly prevalent and the main cause of years lived with disability (1). Consequently, it is a major contributor to health care costs (2). It is defined as ‘‘pain and discomfort’’ localized below the costal margin and above the inferior gluteal folds, with or without leg pain (3). In 85 - 90% of cases, low back pain cannot be attributed to a specific pathology and is hence classified as nonspecific (4).

Arterial tortuosity has been associated with age, elastin deficiency, genetic defects, and cardiovascular risk factors (5–7). It develops due to vascular elongation between two fixed points, forcing the vessel into a tortuous shape (8). While clinically silent in most cases (9), it has been associated with intra-arterial and downstream pathologies, such as dissection, leukoaraiosis, or stroke (10;11).

However, tortuous arteries can also affect their surroundings by thrust. Thrust is a combination of the reaction force owing to flow deflection in the bend, and the blood pressure acting on the end surfaces of the bend. Both forces act in the direction of the bend, thus creating a “pulsatility lens” that enhances pulsatility and focuses it in one direction (Fig. 1). Carotid artery tortuosity has been shown to be associated with vocal cord palsy when targeting the vagus nerve, suggesting pulsatile thrust as the underlying mechanism (13). Imaging in cases like Fig. 2 made me suspect that this mechanism may cause lateral low back pain, hypothesizing that thrust from tortuous iliac arteries inflicts pulsatile microtrauma on the psoas muscle and/or the lumbar nerve roots embedded in it, and that the pain resulting from this interaction is perceived as low back pain.

**Fig. 1.**
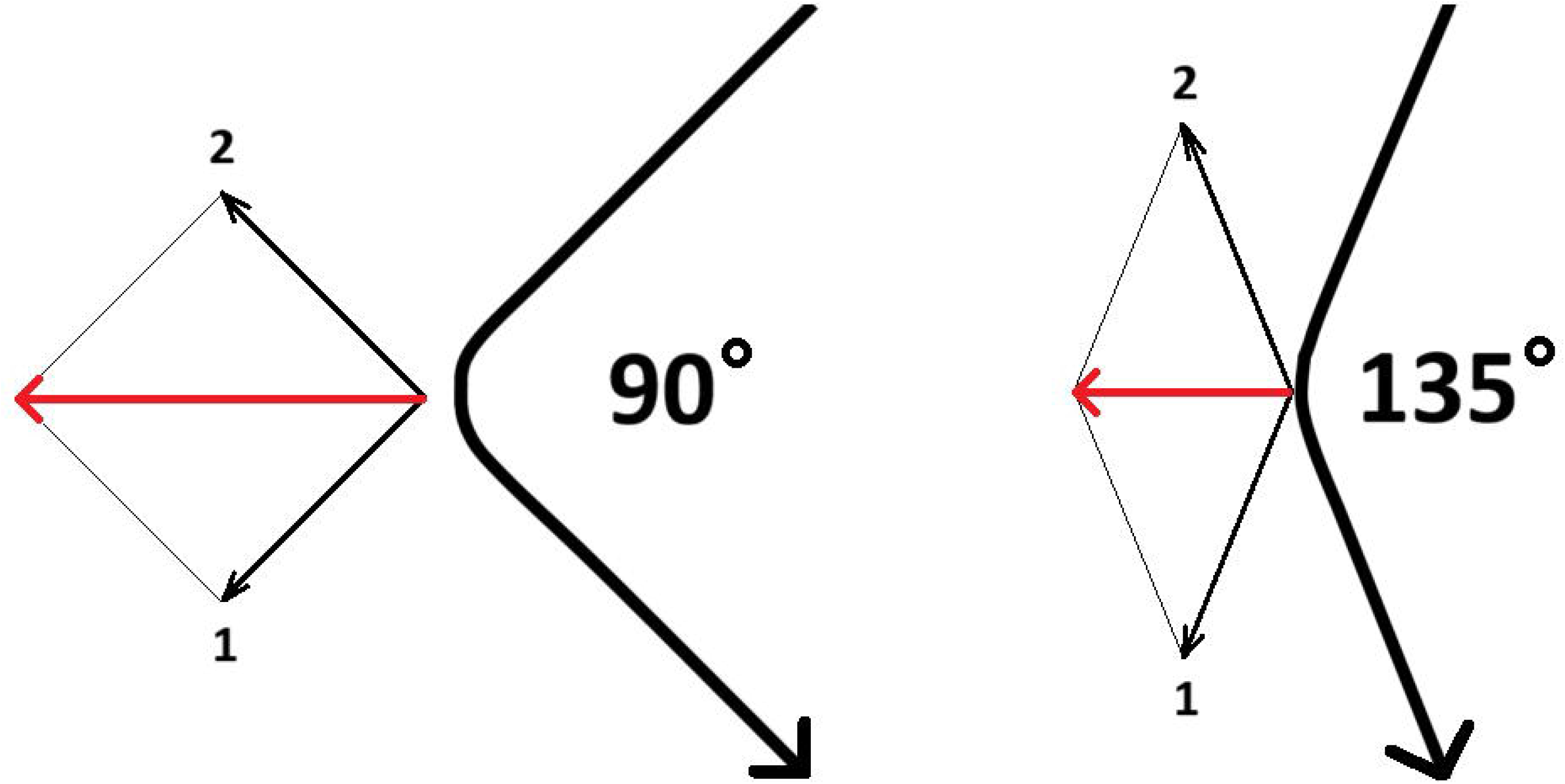
Simplified vector diagram illustrating thrust. Thrust is a trigonometric function depending on the angle of the bend, fluid mass flow, and blood pressure. It acts in the direction of the bend. It is constant in veins and pulsatile in arteries. In an artery with a lumen diameter of 1 cm, a peak systolic flow of 100 cm/s and a systolic blood pressure of 120 mmHg, the systolic thrust caused by flow deviation and blood pressure can be calculated (12) to 1 N in a 135° bend and to 1,9 N in a 90° bend.

**Fig. 2.**
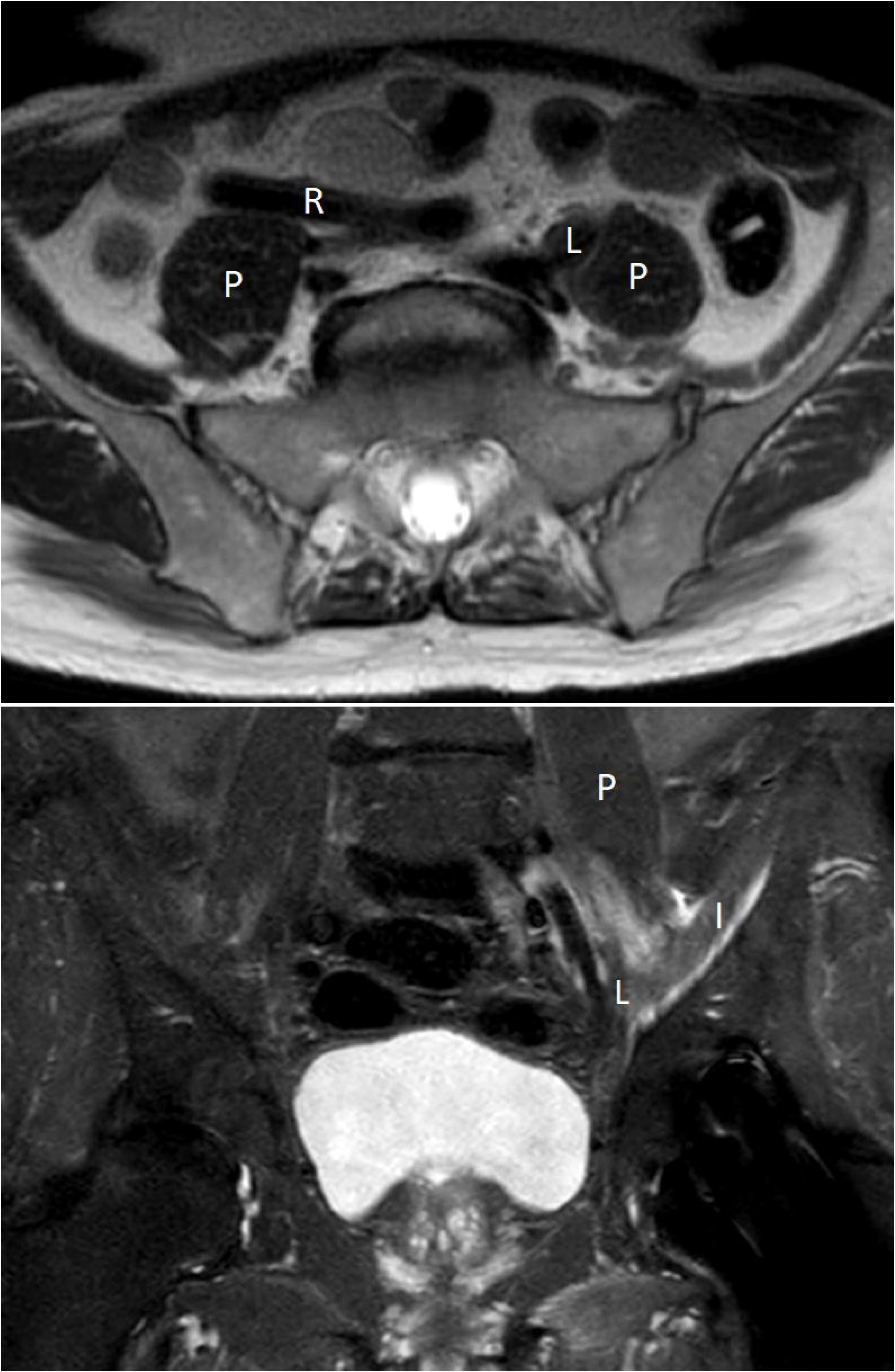
Case report. 67-year-old outpatient referred to magnetic resonance imaging (MRI) of the lumbar spine and pelvis with a four-week history of left sided low lumbar pain irradiating to the front of the thigh, and reduced force on knee extension. MRI of the lumbar spine (not depicted) only showed mild spondylosis. MRI of the pelvis showed marked tortuosity of both common iliac arteries. Top: axial T2 image at level L5/S1 showing the right arterial tortuosity (R) passing in front of the psoas muscle (P), while the left (L) artery’s bend is targeting the psoas muscle, causing an impression in the muscle. Bottom: coronal STIR image showing focal muscular edema (seen as high signal intensity) in the left iliac (I) and psoas (P) muscle at the level of the impression. The angle of this left iliac tortuosity measured on adjacent coronal images (not depicted) was 110°.

If this hypothesis is correct, pain in such cases is presently categorized as nonspecific low back pain, a condition that is not routinely imaged (14). However, iliac artery tortuosity can only be assessed with imaging. Therefore, a retrospective approach based on existing imaging was chosen for this study.

## Material and methods

This retrospective case-control study was performed at the department of radiology at a Danish general hospital between August 2021 and July 2022 by a consultant radiologist with >20 years of experience. The Danish national review board waived the need for approval (NVK 2107163).

### Imaging

All imaging data were obtained from outpatient abdominal computed tomography (CT) scans in the arterial or venous phase after intravenous injection of 100 ml Visipaque 320 (GE Healthcare, Chicago, IL, USA). CT imaging (acquisition thickness, 0.625 mm; pitch, 0.984; 120 kV) was performed on GE Revolution scanners (GE Healthcare, Chicago, IL, USA). Reconstructions in the three standard planes had a thickness of 2.5 mm and a spacing of 2 mm.

### Tortuosity of the iliac artery

Arterial tortuosity is mainly observed in the elderly. Therefore, this study focused on individuals aged 40 - 80 years old. It typically affects the common iliac/external iliac main axis in conjunction. There is no generally accepted definition of iliac artery tortuosity. Due to their retroperitoneal course following the pelvic wall, iliac arteries do not tend to be entirely straight. Therefore, a winding course must be considered normal up to a certain degree. For this study, normal iliac arteries were arbitrarily defined as angles in all iliac artery main axis curvatures of >135° in the coronal and sagittal planes. Marked iliac artery tortuosity was arbitrarily defined as the occurrence of any ≤90° bend in the iliac artery main axis; the 90° angle was chosen because iliac arteries with this angle look tortuous, and because this angle was easy to estimate visually when screening the large numbers of CTs needed for this study. Iliac arteries bending between 90° and 135° were classified as borderline tortuosity and were not included.

The iliac artery main axes were evaluated by examining the standard coronal and sagittal CT reconstructions, without identifying the origin of the internal iliac arteries. No supplementary CT reconstructions were made (Fig. 3).

**Fig. 3.**
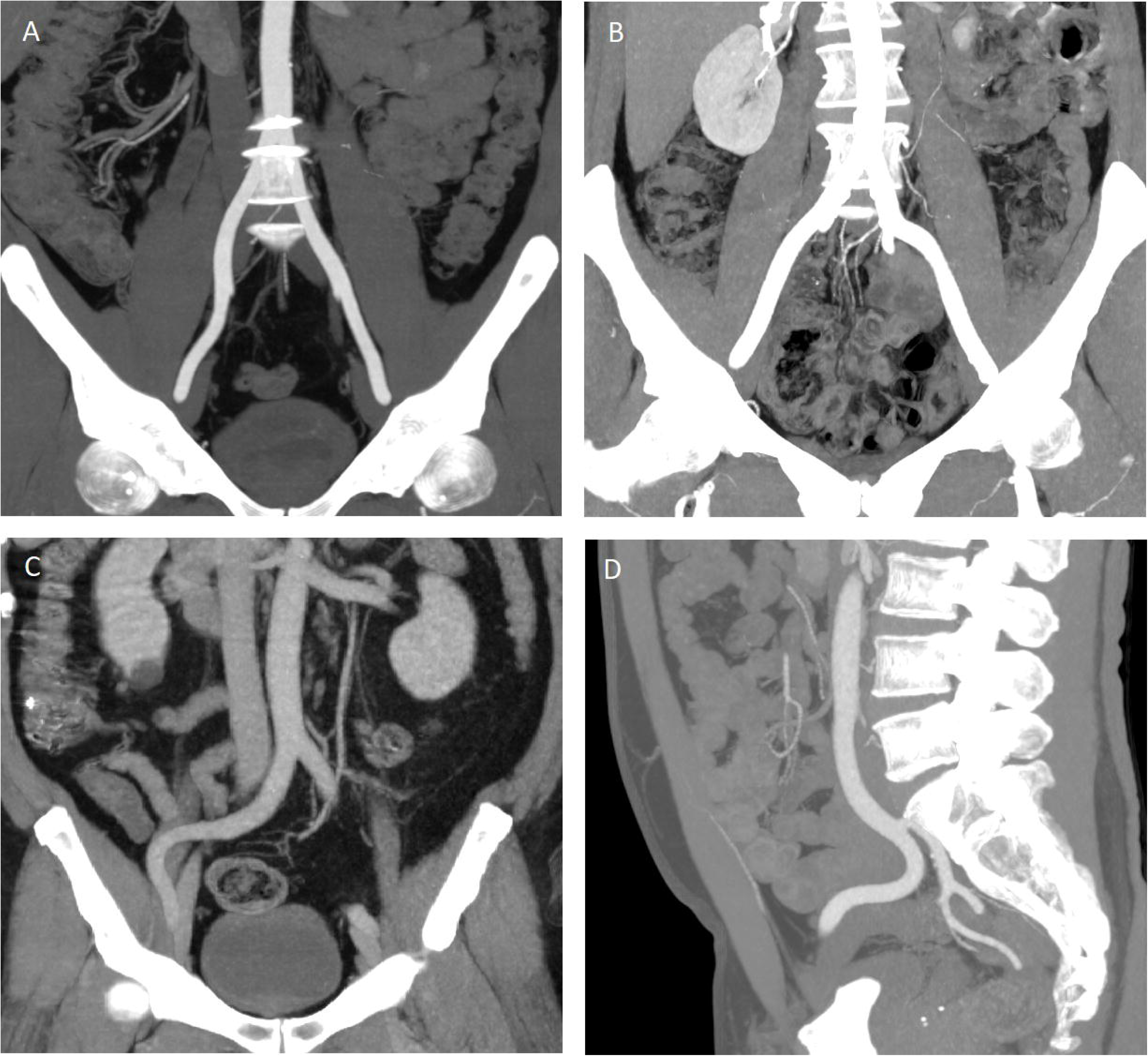
Normal and tortuous iliac arteries. CT maximum intensity projections, thickness 25 mm. A (coronal): normal iliac artery main axes. B (coronal): 135° tortuosity of left iliac artery is classified as borderline normal. C (coronal): marked right sided iliac tortuosity oriented laterally, passing in front of the psoas muscle. D (sagittal): marked right sided iliac tortuosity near midline, oriented posteriorly.

The “pulsatility lens” mechanism can only deliver focused pulsatility to the suspected vulnerable structures (the psoas muscle and/or the lumbar nerve roots embedded in it) in arterial bends directed towards the psoas muscle. Therefore, CT scans of participants with marked iliac tortuosity were reviewed in the axial plane at the level of the peak of the arterial convexity to evaluate whether the tortuosity targets the psoas muscle or passes in front of or behind the muscle (Fig. 4).

**Fig. 4.**
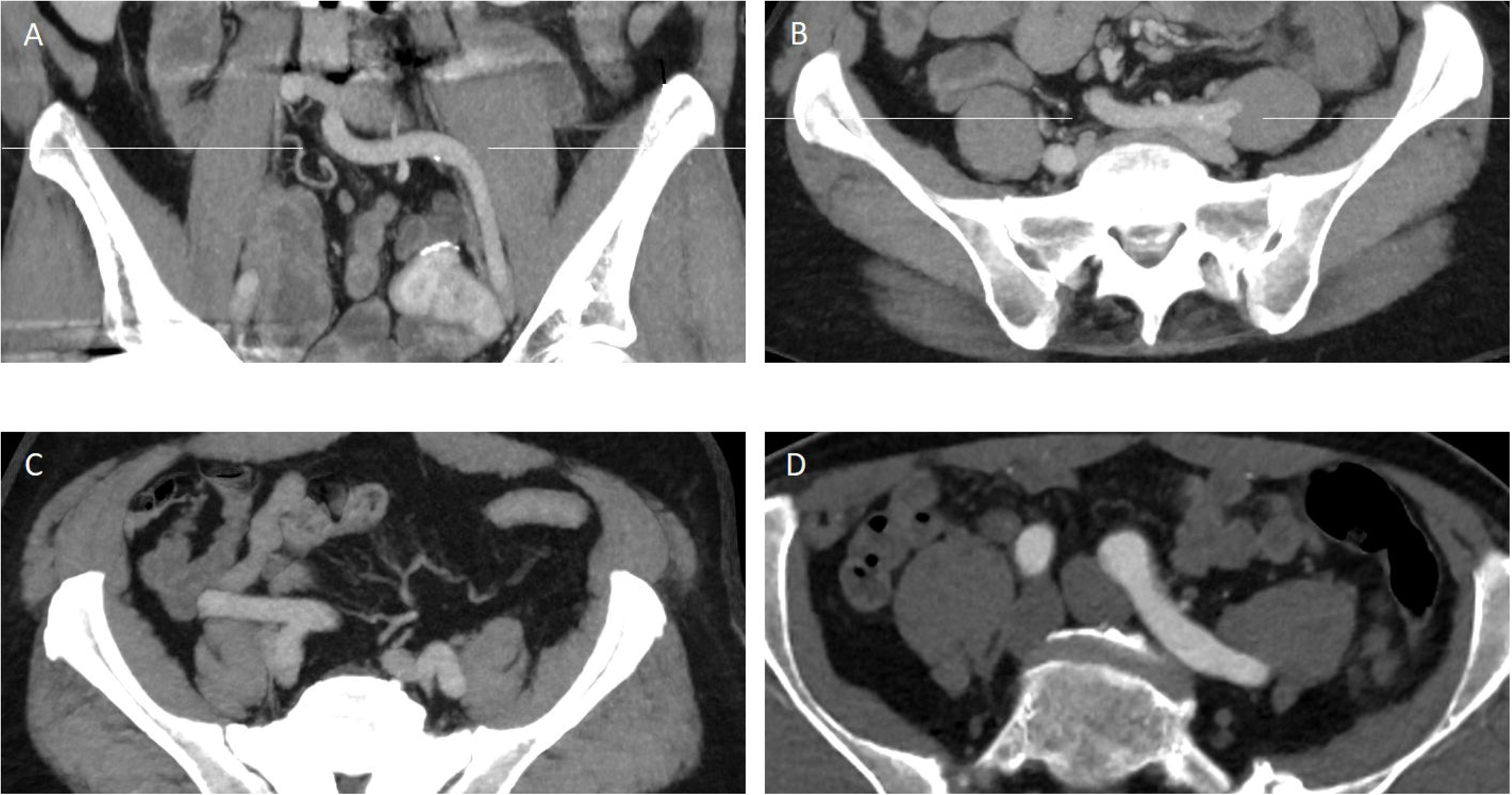
Marked iliac tortuosity and its relation to the psoas muscle. CT maximum intensity projections, thickness 25 mm. A and B are images from the same subject with marked tortuosity of the left iliac artery main axis. The thin white line on A (coronal) indicates the level of axial reconstruction (B), showing the left iliac tortuosity targeting the center of the left psoas muscle. The thin white line on B indicates the coronal reconstruction level (A). C and D are axial images obtained from different subjects. C: Right iliac tortuosity passing in front of the psoas muscle. D: Left iliac tortuosity passing behind the psoas muscle.

### Participants

Participants were identified by continuously screening my department’s outpatient production for abdominal CT scans showing no significant abnormality, selecting individuals with marked tortuosity of the iliac arteries on one or both sides (study group) or normal iliac arteries on both sides (control group). The following exclusion criteria (notion in referral or signs at CT) were applied: malignant disease; degenerative lumbar disease (defined as reduced height of intervertebral discs, herniated disc, neuroforaminal or spinal stenosis); lumbar surgery, fracture, or structural deformity; axial arthritis; aneurisms of the abdominal aorta or iliac arteries; femoral hernia. Within two weeks of CT, subjects qualifying for inclusion were invited by a message sent from the regional health organization using the secure Danish digital platform “e-boks”. After reading the invitation and study description, participants could choose between opting out or giving informed consent and completing a REDCap-based questionnaire (REDCap, Vanderbilt University, TN, USA) for low back pain designed for this study (Table 1). In an attempt to facilitate participation, this questionnaire was kept as simple as possible.

**Table 1.** Questionnaire.

|  |  |
| --- | --- |
| **Do you suffer from low back pain?** | never - occasionally - often - constantly |
| **If yes, is the intensity of pain** | light - moderate - severe |
| **Where is your low back pain localized?** | right - central – left* |
| **Have you suffered from low back pain during the last two weeks?** | yes - no |
| **Do you have pain irradiating to the thigh?** | never - occasionally - often - constantly |
| **If yes, is the intensity of pain** | light - moderate - severe |
| **Localization of irradiating pain:** | front right - back right - front left - back left* |
| **Did you have pain irradiating to the thigh during the last two weeks?** | yes – no |

\*: more than one answer could be given

Concomitantly with the screening of outpatient CT scans for participants in the main study, I reviewed 500 consecutive outpatient CT scans in the age group 40 - 80 years for presence of marked iliac tortuosity without applying any exclusion criteria.

### Sample size and power considerations

This is a first study. It was conceived without existing data about the frequency of lateral low back pain and iliac tortuosity in the background population. Using Bowers’ method and hypothetical values for p1=0.4 (probability of a complaint of lateral low back pain if a tortuous iliac artery targets the psoas muscle), p2=0.2 (probability of a complaint of lateral low back pain in a patient without iliac tortuosity targeting the psoas muscle), and a significance level of p<0.05 gave an estimated sample size of n=80.

### Statistics

The iliac arteries and psoas muscles are paired organs. An affection of the psoas muscle on one side was hypothesized to cause ipsilateral symptoms. The occurrence of this phenomenon on both sides was considered possible. The participant’s right and left sides were regarded as independent entities. Calculations were made for individuals and for sides.

The age distribution of the study and control groups was compared using Student’s t-test. All remaining comparisons were performed on 2 x 2 contingency tables analyzed with the chi-square test, alternatively Fisher’s exact test for discrete variables in those comparisons where one or more of the variables were <5. All analyses were two sided. The level of statistical significance was set at p<0.05. Chi-square and Fisher’s exact test were performed on https://medcalc.org. The remaining statistics were performed on spreadsheet software (Microsoft Excel).

## Results

### Frequency of marked iliac tortuosity

The frequency of marked iliac artery tortuosity on one or both sides in the analysis of 500 consecutive CT scans is shown in Table 2. The tortuosity frequency increased with age. In individuals >70 years, marked tortuosity on one or both sides was found in >50%. Overall, marked iliac tortuosity was more frequent in males (p<0.001).

**Table 2.** Frequency of marked iliac artery tortuosity in 500 consecutive outpatient CT scans.

|  | **40 - 50 y** | **50 - 60 y** | **60 - 70 y** | **70 - 80 y** | **40 - 80 y** |
| --- | --- | --- | --- | --- | --- |
| **total [n] (m; f)** | 24; 27 | 75; 78 | 69; 67 | 76; 84 | 244; 256 |
| **tortuous [n] (m; f)** | 7; 3 | 30; 19 | 37; 15 | 46; 39 | 120; 76 |
| **tortuous [%] (m; f)** | 29; 11 | 40; 24 | 54; 22 | 60; 46 | 49; 30 |
| **tortuous [%] total** | 19.6 | 32 | 38.2 | 53.1 | 39.2 |

### Study population and analysis based on individuals

A total of 304 individuals qualified for inclusion in the study and were invited to participate. A total of 99 participants accepted and returned the questionnaire. Most participants were referred for CT by their general practitioner for suspicion of malignant disease or nonspecific abdominal symptoms. A minor proportion of participants was scanned for follow-up of an aneurism of the thoracic aorta. All questionnaires were returned within four weeks from CT. 52 of the participants had normal iliac arteries (control group), 47 had marked iliac tortuosity on one or both sides (study group). 27 of the participants in the study group had bilateral marked iliac tortuosity, and 18 (2) had unilateral marked tortuosity on the left (right) side; marked iliac tortuosity was more frequent on the left (p<0.001). All iliac tortuosities targeting the psoas muscle did so on the same side.

Significant differences in age (p<0.001) and sex (p<0.001) distribution were observed. To control for confounding from these differences, all comparisons between study group and control group were based on subgroups consisting of matched pairs (17 male and 14 female pairs; age difference between paired subjects ≤2 years; no significant difference in age between the matched groups).

Table 3 shows the demographic data and questionnaire results of the original and matched groups. Lateral low back pain on one or both sides was reported more often by participants in the study group (n=14) than in the control group (n=6), p=0.031.

**Table 3.**
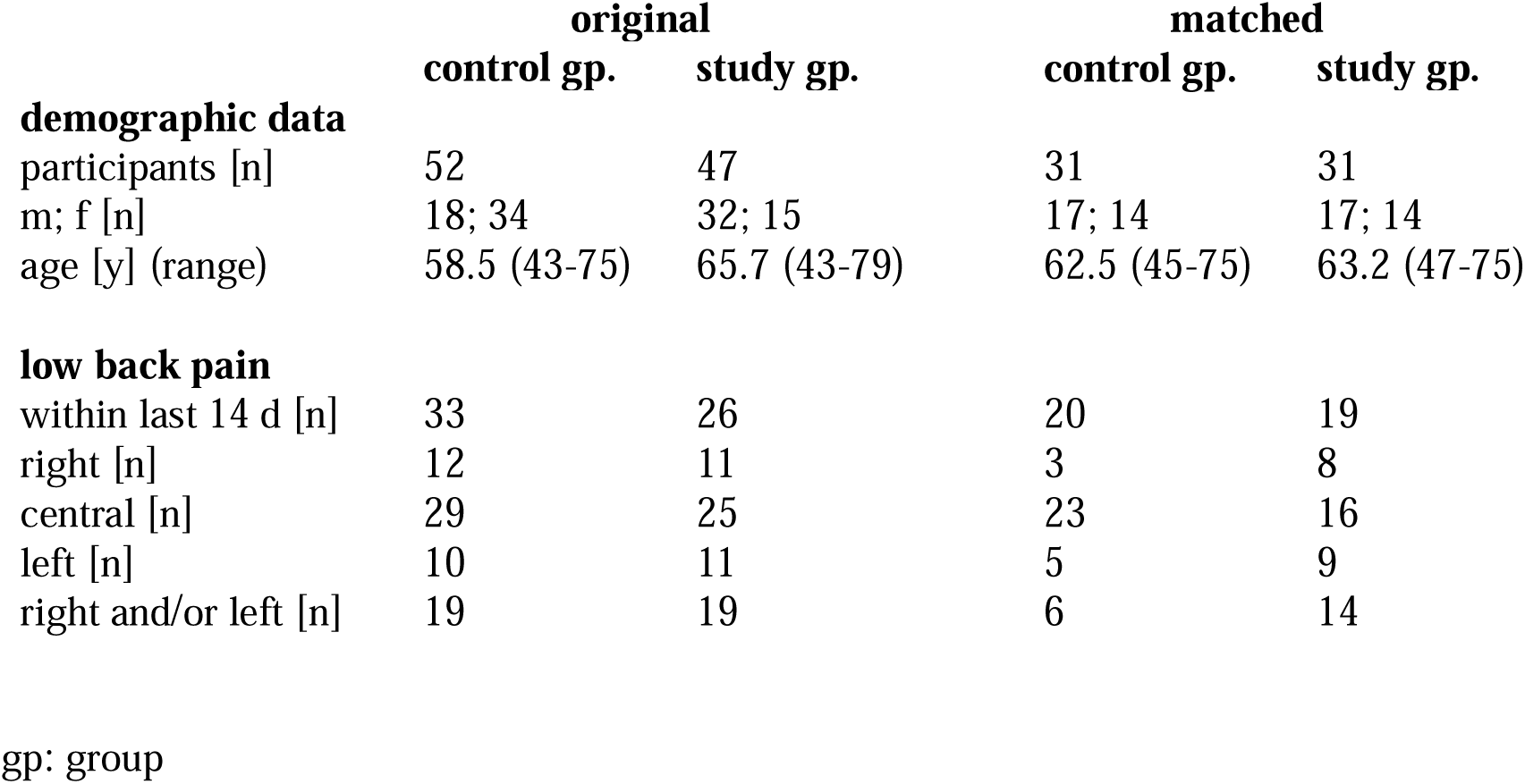
Demographic data and low back pain questionnaire results in the original groups and the matched groups.

|  | **original** |  | **matched** |  |
| --- | --- | --- | --- | --- |
|  | **control gp.** | **study gp.** | **control gp.** | **study gp.** |
| **demographic data** |  |  |  |  |
| participants [n] | 52 | 47 | 31 | 31 |
| m; f [n] | 18; 34 | 32; 15 | 17; 14 | 17; 14 |
| age [y] (range) | 58.5 (43-75) | 65.7 (43-79) | 62.5 (45-75) | 63.2 (47-75) |
| **low back pain** |  |  |  |  |
| within last 14 d [n] | 33 | 26 | 20 | 19 |
| right [n] | 12 | 11 | 3 | 8 |
| central [n] | 29 | 25 | 23 | 16 |
| left [n] | 10 | 11 | 5 | 9 |
| right and/or left [n] | 19 | 19 | 6 | 14 |

gp: group

### Analysis based on sides

Hypothesizing that affection of the psoas muscle on one side causes symptoms on the same side, calculations were also performed for sides (31 subjects = 62 sides). These results are shown in Table 4. Comparing the 62 sides of the control group with all 62 sides of the study group showed significantly more complaints of lateral low back pain in the study group with a p=0.048. However, the iliac arteries were not markedly tortuous on 15 of the 62 sides in the study group. Therefore, an additional comparison was made, including only the 47 sides with marked tortuosity and resulting in a p=0.019. Finally, only the 20 sides with marked tortuosities targeting the psoas muscle were compared with the 62 sides in the control group, resulting in a p=0.0003. The odds ratio for a complaint of lateral low back pain is considerably higher in markedly tortuous iliac arteries targeting the psoas muscle.

**Table 4.** Sides-based comparisons of the frequency of lateral low back pain.

|  | control group |  | study group |  |  |  |  |  |
| --- | --- | --- | --- | --- | --- | --- | --- | --- |
|  | all sides |  | all sides |  | tortuous sides |  | targeting psoas |  |
| sides [n] | 62 | 31R+31L | 62 | 31R+31L | 47 | 18R+29L | 20 | 3R+17L |
| pain [n] | 8 | 3R+5L | 17 | 8R+9L | 15 | 7R+8L | 11 | 3R+8L |
| P |  |  | 0.048 |  | 0.019 |  | 0.0003 |  |
| OR |  |  | 2.55 |  | 3.16 |  | 8.25 |  |
| OR 95% CI |  |  | 1.01 to 6.46 |  | 1.2 to 8.29 |  | 2.6 to 26.11 |  |

R: right; L: left; OR: odds ratio; CI: confidence interval

The sidedness approach also allowed for comparison of both sides within the same subject, obviating the comparison with the control group and thus avoiding the confounding related to these comparisons. Therefore, participants for this comparison were recruited from the original study group (n=47). A total of 21 participants in this group had marked iliac tortuosity targeting the psoas muscle on one side, and a normal artery or a tortuosity *not* targeting the psoas on the other (10f with mean age 66.3y + 11m with mean age 63.7y). In 20 of these 21 subjects, the psoas was targeted on the left side. Lateral pain on one or both sides was reported by 11 of these subjects (mean age 66.8y) and no lateral pain by 10 subjects (mean age 62.9y). Lateral pain was reported on 9 of 21 sides with iliac tortuosities targeting the psoas muscle, but only on 2 of 21 sides without targeting of the psoas muscle: the side of lateral pain correlates with the side of a tortuous iliac artery targeting the psoas muscle (p=0.032).

### Other results

The analyses of the participants’ intensity and frequency of low back pain and of symptoms in the lower extremities did not reveal any significant findings – these data are not shown. 9 of the 31 participants in the study group had marked iliac artery tortuosity targeting the psoas muscle on one or both sides and reported lateral low back pain. Marked iliac tortuosity being found in 39,2 % of subjects aged 40 - 80 years, the estimated prevalence of lateral low back pain associated with iliac tortuosity in this age group is 11.4 %.

## Discussion

The findings of my study suggest that tortuous iliac arteries can cause pain by affecting the psoas muscle, and that the pain resulting from this interaction is perceived as lateral low back pain. This provides further evidence of the pathophysiological significance of thrust if the bend of a tortuous artery targets a vulnerable structure.

A related mechanism is found in intracranial neurovascular compression syndromes. Key features for the diagnosis of these syndromes are the localization of the contact between the vessel and cranial nerve at the transition zone between the central and peripheral myelin, and the displacement and atrophy of the affected nerve (15). However, thrust has implicitly been associated with the pathogenesis of neurovascular compression syndromes, as reflected by the suggestion that vessel contact should be perpendicular to the axis of the affected nerve (16). Thus, in a series of seven trigeminal neuralgias (17), all causative vessels performed loops oriented towards the nerve. In neurovascular compression (18) as well as in the present study, a significant number of vessel juxtapositions is found in asymptomatic patients. This suggests that hemodynamic phenomena alone may not be sufficient to account for the observed symptoms (19).

This study’s untraditional selection of participants was chosen in order to categorize the iliac arteries before inclusion. The participants in this study can be regarded as a sample from the background population obtained by a combination of existing imaging, exclusion criteria, and the willingness to participate. A considerable proportion of subjects with low back pain can be expected in any sample of the background population: in a review of the worldwide prevalence of low back pain including 165 studies from 54 countries, the 1-month prevalence was 30.8% (20). The even higher (55 to 64.5 %) prevalence of low back pain in my study is probably due to response bias. However, response bias cannot have affected my findings because all participants were ignorant of their vascular status.

Despite its small sample size, my study produces significant results by taking advantage of the bilaterality of the involved structures (psoas muscle and iliac artery), allowing an analysis of the body halves, and producing two independent types of evidence: 1. a higher frequency of lateral low back pain in the study group than in the control group and 2. a within-person correlation that is reminiscent of split-body trials where inter-person variability is removed by making the comparison within the same person (21). The exclusion of borderline iliac tortuosity (angles between 90° and 135°) secured a significant difference in iliac pulsatility between the control group and the study group. Thus, the estimated 11.4 % prevalence of the described association in subjects aged 40 - 80 years may be a conservative estimate because the association is also likely to be found in many cases of borderline iliac tortuosity (the causative vessel in Fig. 2 had an angle of 110°) that was not included in this study.

In conclusion, my study found an association between tortuosity of the iliac artery targeting the psoas muscle and ipsilateral low back pain. This association may explain lateral low back pain in part of the cases currently regarded as nonspecific. Demarcating lateral low back pain associated with iliac tortuosity against nonspecific low back pain may lead to a more specific treatment.

## Data Availability

All data produced in the present study are available upon reasonable request to the author

## Notes

### Competing Interest Statement

The authors have declared no competing interest.

### Author Declarations

The Danish National Review Board waived ethical approval of this work (NVK 2107163).

### Summary of Updates

1. a calculation of the estimated thrust force acting in an iliac bend was added (legend Fig. 1) 2. an calculation of the correlation between side of pain and side of iliac bend targeting the psoas muscle in participants of the study gropu was added, obviating the comparison with the control group and thus avoiding the confounding related to these comparisons.

